# Safety of Repeated Low-Level Red-Light Therapy for Myopia: A Systematic Review

**DOI:** 10.1101/2024.04.19.24306057

**Authors:** Yanping Chen, Shida Chen, Ruilin Xiong, Shaopeng Yang, Riqian Liu, Ziyu Zhu, Kaidi Xiang, Nathan Congdon, Wei Wang

## Abstract

**Topic:** Existing evidence for the safety of repeated low-level red-light (RLRL) therapy for myopia control.

**Clinical relevance:** Recent trials show RLRL therapy is effective in the prevention and control of myopia. Establishing its safety profile is necessary prior to widespread clinical implementation.

**Methods:** We conducted a systematic review (International Prospective Register of Systematic Reviews, CRD42024516676) of articles across seven databases from inception through February 10, 2024, with keywords related to myopia and RLRL therapy. Pooled safety outcomes and risk-to-benefit ratios were reported, and incidence of side effects was compared with other anti-myopia interventions. Quality appraisal was performed using the Cochrane Risk of Bias Tool.

**Results:** Among 689 screened articles, 20 studies (2.90%; eleven randomized controlled trials, four non-randomized controlled trials, one post-trial study, one single-arm study, one retrospective study and two case reports of identical patient.; median duration 9 months, longest 24 months) were analysed, encompassing 2,380 participants aged 3-18 years and 1,436 individuals undergoing RLRL therapy. Two case reports described an identical patient with reversible decline in visual acuity and optical coherence tomography (OCT) abnormalities, completely resolved 4 months after treatment cessation. No cases of permanent vision loss were reported. Temporary afterimage was the most common ocular symptom following treatment, resolving within 6 minutes in reported studies. The number needed to harm outweighed the number needed to treat by a ratio of 12.7-21.4 for a person with −3D to −8D myopia treated with RLRL therapy. Incidence of side effects from RLRL was 0.088 per 100 patient-years (95% confidence interval [CI], 0.02-0.50), comparable to spectacles designed for myopia reduction (0.22; 95% CI, 0.09-0.51; P=0.385), and significantly lower than for low-dose atropine (7.32; 95% CI, 6.65-8.05; P<0.001), orthokeratology (20.6; 95% CI, 16.7-25.0; P<0.001), other anti-myopia contact lens (19.3; 95% CI, 17.6-21.1; P<0.001).

**Conclusion:** No irreversible visual function loss or ocular structural damage was identified with RLRL. Fundus photography and OCT before and during therapy, alongside home monitoring of visual acuity and duration of afterimages, are necessary to identify side effects. Further adequately-powered studies of longer duration are needed to evaluate long-term safety of RLRL.

## Introduction

Myopia is the most common ocular disorder of childhood and adolescence. Without effective intervention measures, it is estimated that approximately 50% of the global population will be affected by 2050.^1^ The rising prevalence of myopia, along with earlier onset, increases the risk of high myopia, which may be associated with irreversible, sight-threatening complications such as myopic maculopathy, glaucoma and retinal detachment.^2–4^ Therefore, prevention and control of myopia have become important public health challenges.

Repeated low-level red-light (RLRL) therapy, highlighted in the latest 2023 Digest of the International Myopia Institute (IMI), has emerged as a novel approach to myopia prevention.^5^ RLRL therapy involves locally irradiating the retina with low-level red light (approximately 1600 lux). It falls under Group 1 of the ANSI Z80.36-2021 standard,^6^ ensuring its safety for clinical ophthalmic applications. Unlike traditional therapeutic lasers such as panretinal photocoagulation, which rely on thermal effects generated through transpupillary energy ranging from 200-250 mW passing through the pupil, RLRL uses a much lower energy level of 0.29 mW, avoiding any thermal effects. Its therapeutic impact is postulated to rely on photobiomodulation (PBM) secondary to the laser’s energy, potentially leading to thickening of the choroidal layer.^7^

Growing trial evidence supports the effectiveness of RLRL therapy in reducing myopia progression. The first multicenter randomized controlled trial (RCT) in China demonstrated in 2019 that home-based RLRL therapy, administered for 3 minutes twice daily for 5 days a week, significantly reduced axial elongation by 69.4% and refractive progression by 76.6% among school-aged children.^8^ Subsequent clinical trials have consistently confirmed these findings.^9–11^ Recently, a meta-analysis incorporating 13 studies comprising 8 RCTs, 3 non-randomized controlled trials and 2 cohort studies, and involving a total of 1857 children and adolescents, confirmed the efficacy of RLRL therapy.^12^ Six-month weighted differences in spherical equivalent refraction (SER) and axial length (AL) were 0.68 diopters (D) and 0.35 mm respectively between the RLRL treatment group and controls. However, published studies have predominantly focused on efficacy outcomes, with limited reporting of the safety and side effects of RLRL therapy.

To address this important knowledge gap, we systematically reviewed the safety profiles and risk-to-benefit ratio associated with RLRL treatment for myopia prevention and control. The aim of this study is to provide insights to help both clinicians and program designers optimize use of RLRL therapy, ensuring its safe and effective integration into myopia management.

## Methods

This review was registered prospectively on the International Prospective Register of Systematic Reviews database (available from https://www.crd.york.ac.uk/prospero/display_record.php?ID=CRD42024516676) and is reported in accordance with the Preferred Reporting Items for Systematic Reviews and Meta-Analyses (PRISMA) statement.^13^ All research adhered to the tenets of the Declaration of Helsinki. Individual patient-level consent was not required, nor was ethical review.

### Eligibility Criteria

We included clinical studies of RLRL therapy designed to prevent myopia or delay its progression. Studies were selected according to the following criteria: (1) Participants were younger than 18 years with myopia or premyopia; (2) RLRL therapy was used in at least one arm or by all participants; (3) Reporting of at least 1 safety outcome, including visual function, ocular structural assessment, or adverse events; (4) All study types except literature reviews, in order to capture as many reported safety profiles and side effects as possible. We excluded studies on non-human subjects, those enrolling participants with secondary myopia, without safety data, and those merely describing treatments combined with RLRL therapy or involving red light flicker rather than continuous administration.

### Search Strategy

A comprehensive search of the peer-reviewed literature was conducted across seven databases, including PubMed, Embase, Web of Science, Cochrane Library, Scopus, the Chinese databases China National Knowledge Infrastructure and VIP Information Database, from their dates of inception through 10 February 2024. We used a combination of key words related to myopia and RLRL therapy, imposing no language restrictions. To ensure thorough coverage, we also scrutinized relevant reviews and all references cited by eligible studies for additional pertinent publications. The full search strategy is detailed in **Table S1**.

### Study Selection

Citations retrieved from electronic databases were compiled into an EndNote library by one author (Y.C.). After the removal of duplicates, two reviewers (Y.C., R.X.) independently screened titles and abstracts to assess initial eligibility. Reviewers then checked the full text for potentially eligible studies to determine their final inclusion or exclusion. The primary reason for exclusion was documented at the full text screening stage. Any discrepancies between reviewers were resolved through discussion or by consulting a third researcher if necessary (W.W.).

### Data Collection and Risk of Bias Assessment

Data were extracted independently by two reviewers (Y.C., R.X.) and were entered into an Excel spreadsheet (version 2022, Microsoft Corporation, Redmond, USA). For each included study, the extracted information consisted of author’s name, year of publication, study design, country or area, specification of the red light device (device name, manufacturer, wavelength and power), treatment regimen, sample size, follow-up duration, age, sex, baseline SER, baseline AL, safety outcomes, and participation completion rate. For the safety outcomes, we documented all safety data reported throughout the selected studies. Two reviewers (Y.C., S.Y.) independently appraised articles for systematic bias using the Cochrane Risk of Bias Tool.^14^

### Data Synthesis and Analysis

Given the heterogeneous reporting of safety outcomes among the included studies, it was not feasible to conduct a pooled meta-analysis.^15^ Instead, we performed a risk-to-benefit analysis according to the model described by Bullimore to assess whether the potential benefits of reducing myopia progression of 1D with RLRL therapy outweigh the potential risks associated with the treatment.^16^ The revised Bullimore’s model is based on three assumptions: 1) Every patient undergoing myopia control will use RLRL for a 5-year treatment period. Since there’s lack of long-term efficacy of RLRL therapy, the selection of five years facilitates the conservative estimation of the effect of controlling myopia progression within 1D as per previous review.^17^ 2) Serious adverse events, if they occur, may happen at a mean age of 12 years during this five-year treatment period.^16^ 3) Based on the estimated mean life expectancy of 77 years in China (https://data.who.int), any adverse event resulting in immediate visual impairment will lead to visual impairment for a duration of 65 years. The annual incidence rate of vision loss was calculated using the number of children experiencing visual impairment from RLRL as the numerator and the estimated total number of patient-years in reported clinical RLRL studies with safety outcomes as the denominator. The absolute risk increase (ARI) of vision loss years was estimated by multiplying the annual incidence of vision loss per 10,000 patients by the assumed duration of visual impairment, which is 65 years. The number needed to harm (NNH) was then calculated as the reciprocal of ARI. This metric indicates the number of patients who need to be treated to induce one case of visual impairment.

According to Bullimore’s model, the benefit of preventing visual impairment from blinding myopic related complications by 1D was regardless of the treatment. The average duration of visual impairment that a patient is likely to experience over their lifetime at myopia ranging from −3D to −8D was estimated. ^16^ For instance, a patient with −3D myopia is expected to experience an average of 4.42 years of visual impairment (mild visual impairment as US definition of 20/40) while a −4D person will experience 5.25 years of visual impairment. Thus, the benefit of slowing myopia progression by 1D is quantified by the difference in years of visual impairment, amounting to a prevention of 0.84 years of visual impairment if myopia control interventions in an individual potentially reaching −4D myopia result in achieving only −3D. The values of years of visual impairment prevented by 1D reduction ranges from 0.74 to 1.22 for −3D to −8D. The number needed to treat (NNT) was evaluated in Bullimore’s model to prevent 1-year (NNT range, 0.82-1.38) and 5-year (NNT range, 4.11-6.75) visual impairment.^16^ Furtherly, we calculated the NNH/NNT ratio for RLRL therapy in patients with myopia degrees ranging from −3D to −8D.

Additionally, we conducted a systematic comparison of the incidence of side effects between RLRL therapy and other anti-myopia interventions. Our approach involved a systematic search of all eligible peer-reviewed RCTs that included myopic or premyopic participants younger than 18 years, had at least a one-year follow-up period, and investigated interventions including low-dose atropine, orthokeratology, other contact lenses, and spectacles. For spectacles, we included bifocal lens, progressive addition spectacles, aspherical lenslets and peripheral defocus spectacles as spectacles designed for myopia reduction since their incidence rates of adverse events were similar. Adverse events of other interventions were counted as the number of events reported in published RCTs. The crude incidence of adverse events was computed per 100 patient-years of intervention, with 95% confidence intervals (CIs) calculated using the Wilson method.^18, 19^ A two-sided p value <0.05 was defined as statistically significant. All data analyses were performed using Stata (version 17.0, StataCorp, College Station, Texas, USA).

## Results

### Study Selection

Of the 689 references retrieved, 39 full-text articles (5.66%) were reviewed, and 20 studies (2.90%) were identified as eligible for systematic review (**Figure 1**). The primary reason for exclusion during the full-text review process was the absence of reported safety outcomes (n=14, 73.7%).^7, 20–32^

**Figure 1.**
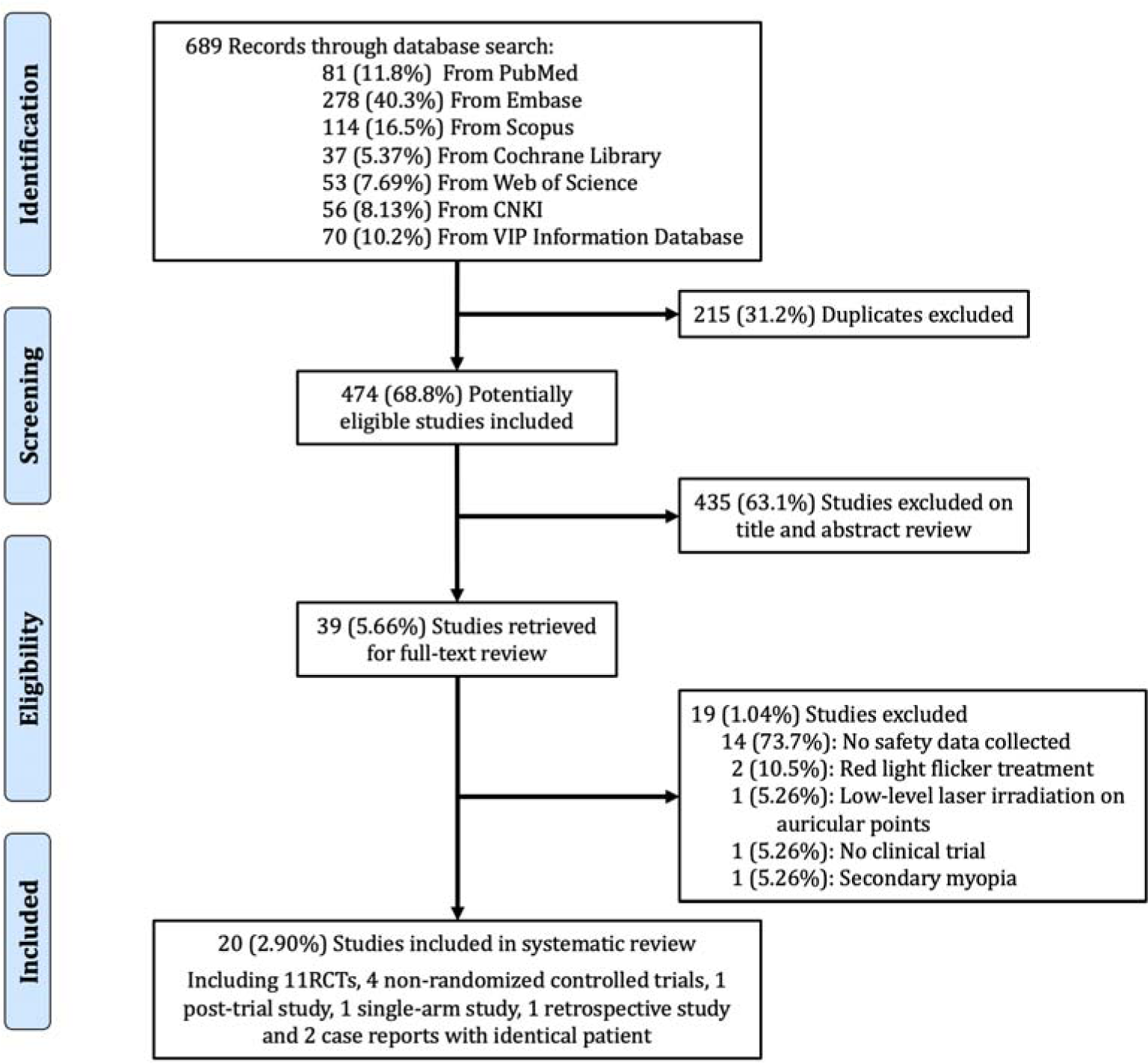
Study selection outlined according to Preferred Reporting Items for Systematic Reviews and Meta-Analyses (PRISMA) guidelines. RCT=randomized controlled trial.

### Characteristics of Included Studies

The 20 studies comprised eleven RCTs,^8, 10, 11, 33–40^ four non-randomized controlled trials,^41–44^ one post-trial study,^9^ one single-arm study,^45^ one retrospective study^46^ and two case reports of identical patient.^47, 48^ Fifteen studies evaluated the safety and efficacy of RLRL therapy compared to controls, eleven with single-vision spectacles (SVS),^8, 10, 33, 35–37, 39–43^ one with a sham device,^11^ one with 0.01% atropine,^34^ and one with orthokeratology, SVS, and combination treatment of orthokeratology and RLRL.^44^ One study compared the safety and efficacy between RLRL devices with different powers and SVS.^38^ Two case reports described the identical individual case with different details;^47, 48^ thus we comprehensively reviewed the two reports but only included as one participant in the systematic review. These studies included a total of 2,380 participants aged 3-18 years presenting at baseline with myopia (cycloplegic SER −0.50 to −9.00D) or premyopia (cycloplegic SER −0.50 to +0.75D), among them 1,436 subjects undergoing RLRL therapy. The median follow-up duration was 9 months, with an interquartile range of 6-12 months, and a longest follow-up period of 24 months. All studies reported a participant completion rate of over 50%, were conducted in China, and published between 2021 and 2024. (**Table 1**). The risk of bias assessment is shown in **Figure 2**.

**Figure 2.**
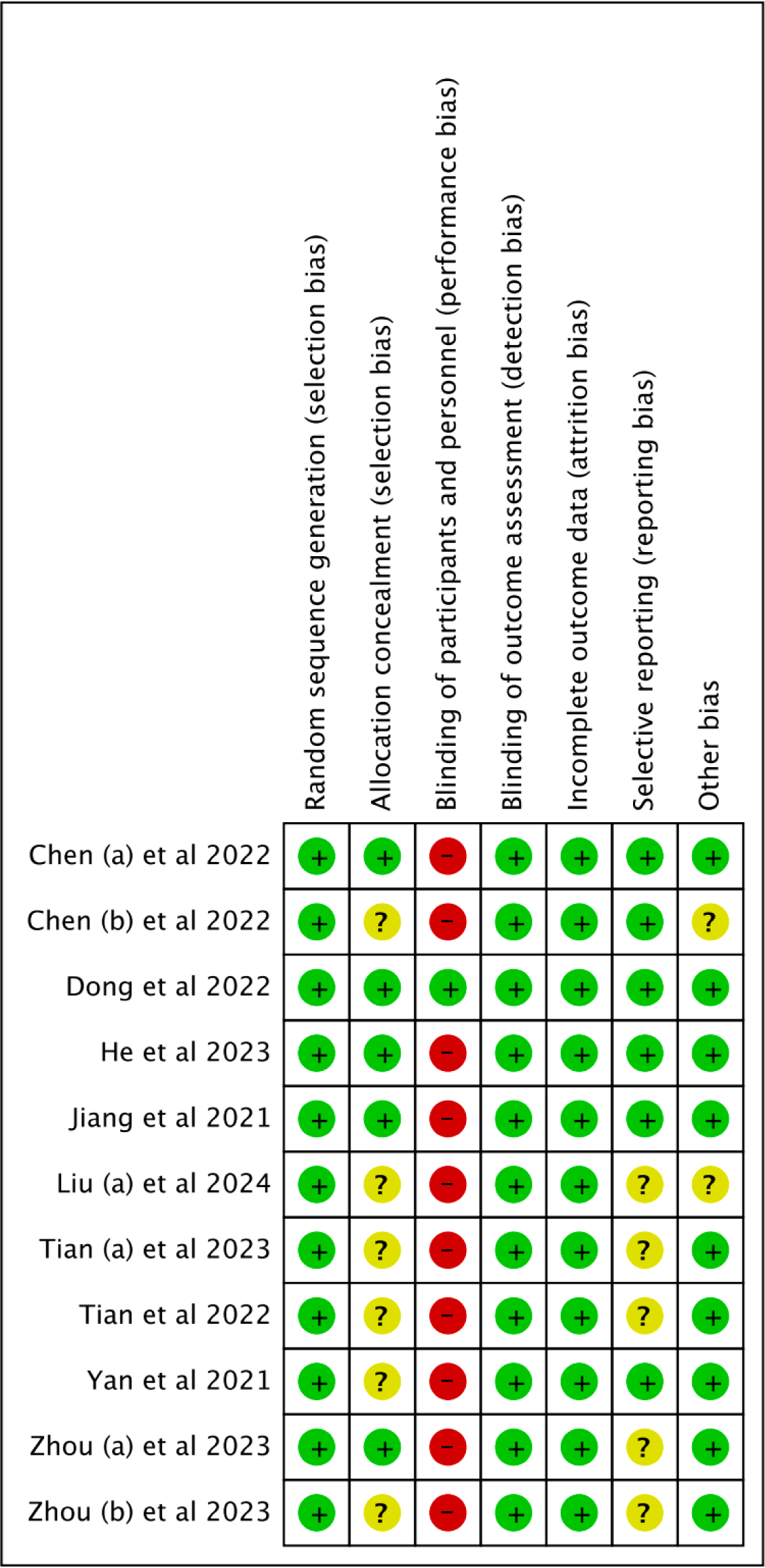
Risk of bias assessment by Cochrane Risk of Bias Tool of the included RCTs. RCT=randomized controlled trial.

**Table 1.** Characteristics of included studies.

| No. of Patients<br>by Study<br>(Study Type) | Treatment | Age (y) | Female (%) | Mean SER (D) | Mean AL<br>(mm) | Completion<br>Rate (%) |
| --- | --- | --- | --- | --- | --- | --- |
| Jiang et al 2021 (RCT)<sup>8</sup> |  |  |  |  |  |  |
| 117 | Intervention (RLRL) | 8-13 | 62 (52.1) | -2.49 (0.92) | 24.5 (0.67) | 98.3 |
| 129 | Control (SVS) | 8-13 | 72 (49.7) | -2.67 (1.06) | 24.6 (0.86) | 89.0 |
| Yan et al 2021 (RCT)<sup>33</sup> |  |  |  |  |  |  |
| 60 | Intervention (RLRL) | 7-12 | 58 (48.3) | -2.52 (1.15) | 24.2 (0.92) | 100 |
| 60 | Control (SVS) | 7-12 |  | -2.53 (1.15) | 24.4 (0.79) | 100 |
| Xiong et al 2022 (post-trial follow-up study)<sup>9</sup> |  |  |  |  |  |  |
| 11 | RLRL-RLRL | 8-13 | 4 (36.4) | -1.77 (0.57) | 24.9 (0.94) | - |
| 10 | SVS-RLRL | 8-13 | 5 (50.0) | -2.57 (1.11) | 24.8 (0.71) | - |
| 52 | RLRL-SVS | 8-13 | 26 (50.0) | -2.50 (0.83) | 24.5 (0.58) | - |
| 41 | SVS-SVS | 8-13 | 26 (63.4) | -2.76 (1.15) | 24.6 (0.94) | - |
| Dong et al 2022 (RCT)<sup>11</sup> |  |  |  |  |  |  |
| 56 | Intervention (RLRL) | 7-12 | 30 (53.6) | -3.13 (1.91) | 24.7 (1.04) | 94.6 |
| 55 | Control (Sham device) | 7-12 | 26 (46.4) | -2.82 (1.86) | 24.6 (0.96) | 98.2 |
| Chen (a) et al 2022 (RCT)<sup>34</sup> |  |  |  |  |  |  |
| 30 | Intervention (RLRL) | 7-15 | 17 (54.8) | -2.60 (1.17) | 24.5 (0.79) | 96.8 |
| 30 | Control (0.01% atropine) | 7-15 | 14 (45.2) | -2.59 (1.24) | 24.7 (0.98) | 96.8 |
| Chen (b) et al 2022 (RCT)<sup>35</sup> |  |  |  |  |  |  |
| 46 | Intervention (RLRL) | 6-13 | 19 (41.3) | -2.54 (1.04) | 24.6 (0.97) | 90.2 |
| 40 | Control (SVS) | 6-13 | 15 (37.5) | -2.29 (0.77) | 24.6 (0.76) | 78.4 |
| Tian et al 2022 (RCT)<sup>36</sup> |  |  |  |  |  |  |
| 91 | Intervention (RLRL) | 6-12 | 55 (49.1) | -2.00 (0.33) | 24.3 (0.92) | 81.3 |
| 88 | Control (SVS) | 6-12 | 57 (50.9) | -2.00 (0.25) | 24.2 (0.85) | 78.6 |
| He et al 2023 (RCT)<sup>10</sup> |  |  |  |  |  |  |
| 126 | Intervention (RLRL) | 6-11 | 68 (48.9) | 0.14 (0.30) | 23.4 (0.68) | 90.7 |
| 122 | Control (SVS) | 6-11 | 71 (51.1) | 0.16 (0.28) | 23.3 (0.69) | 87.8 |
| Zhou (a) et al 2023 (RCT)<sup>37</sup> |  |  |  |  |  |  |
| 20 | Intervention (RLRL) | 3-16 | 9 (37.5) | -2.93 (1.87) | 24.6 (1.16) | 96.0 |
| 15 | Control (SVS) | 3-16 | 7 (41.2) | -2.11 (1.21) | 24.4 (0.87) | 76.0 |
| Zhou (b) et al 2023 (RCT)<sup>38</sup> |  |  |  |  |  |  |
| 43 | Intervention (RLRL 0.37mW) | 6-15 | 20 (40.0) | -1.72 (0.91) | 24.2 (0.79) | 86.0 |
| 47 | Intervention (RLRL 0.60mW) | 6-15 | 25 (50.0) | -2.01 (0.87) | 24.1 (0.88) | 94.0 |
| 44 | Intervention (RLRL 1.20mW) | 6-15 | 24 (48.0) | -2.08 (1.33) | 24.5 (0.91) | 88.0 |
| 43 | Control (SVS) | 6-15 | 21 (42.0) | -2.10 (0.90) | 24.4 (0.90) | 86.0 |
| Tian (a) et al 2023 (RCT)<sup>39</sup> |  |  |  |  |  |  |
| 56 | Intervention (RLRL) | 6-12 | 33 (58.9) | 0.25 (0.25) | 23.1 (0.80) | 100 |
| 56 | Control (SVS) | 6-12 | 31 (55.4) | 0.25 (0.19) | 23.1 (0.70) | 100 |
| Lin et al 2023 (non-randomized controlled trial)<sup>41</sup> |  |  |  |  |  |  |
| 41 | Intervention (RLRL) | 6-18 | 84 (51.2) | -3.20 (2.82) | 24.3 (1.04) | 58.6 |
| 58 | Intervention (RLRL) | 6-18 |  | -7.93 (2.95) | 25.7 (1.57) | 82.9 |
| 65 | Control (SVS) | 6-18 |  | -2.32 (2.64) | 24.3 (1.21) | 92.9 |
| Zhao et al 2023 (non-randomized controlled trial)<sup>42</sup> |  |  |  |  |  |  |
| 47 | Intervention (RLRL) | 6-18 | 18 (38.3) | -2.31 (1.26) | 24.6 (0.88) | 100 |
| 20 | Control (SVS) | 6-18 | 8 (40.0) | -2.75 (0.84) | 24.8 (0.90) | 100 |
| Wang et al 2023 (retrospective study)<sup>46</sup> |  |  |  |  |  |  |
| 434 | Intervention (RLRL) | 3-17 | 200 (46.1) | -3.74 (2.60) | 24.9 (1.20) | - |
| Liu et al 2023 & Tian (b) et al 2023 (case report)<sup>47,48</sup> |  |  |  |  |  |  |
| 1 | Intervention (RLRL) | 12 | 1 (100) | OD -6.75D<br>OS -6.25D | - | - |
| Liu (a) et al 2024 (RCT)<sup>40</sup> |  |  |  |  |  |  |
| 47 | Intervention (RLRL) | 7-12 | 19 (44.2) | 0.17 (0.35) | 23.6 (0.78) | 91.5 |
| 47 | Control (SVS) | 7-12 | 20 (47.6) | 0.30 (0.35) | 23.3 (0.73) | 89.4 |
| Liu (b) et al 2024 (single-arm study)<sup>45</sup> |  |  |  |  |  |  |
| 40 | Intervention (RLRL) | 7-14 | 17 (42.5) | -2.75 (1.43) | 24.9 (0.97) | 100 |
| Xiong et al 2024 (non-randomized controlled trial)<sup>43</sup> |  |  |  |  |  |  |
| 45 | Intervention (RLRL) | 7-16 | 22 (48.9) | -3.00 (0.90) | 23.6 (0.35) | 100 |
| 45 | Control (SVS) | 7-16 | 23 (51.1) | -3.02 (0.11) | 23.6 (0.37) | 100 |
| Zhang et al 2024 (non-randomized controlled trial)*<sup>44</sup> |  |  |  |  |  |  |
| 44 | Intervention (RLRL) | ≥7 | 25 (56.8) | -1.87 (1.16) | 24.1 (0.80) | 100 |
| 32 | Intervention (OK) | $\geq 7$ | 14 (43.8) | -2.44 (1.15) | 24.2 (0.73) | 100 |
| 29 | Intervention (RLRL+OK) | $\geq 7$ | 10 (34.5) | -2.55 (1.32) | 24.5 (0.96) | 100 |
| 36 | Control (SVS) | $\geq 7$ | 18 (50.0) | -2.11 (1.11) | 24.3 (0.82) | 100 |

\* Only eye number for each group and total participant number was available in the publication.
Data were presented as number (percentage) or mean (standard deviation).
RLRL=repeated low-level red-light; SVS=single vision spectacles; RCT=randomized controlled trial; SER= spherical equivalent refraction; AL=axial length; OK=orthokeratology.

The specifications of RLRL therapy and treatment regimens across the included studies are summarized in **Table S2**. Four types of RLRL devices were utilized, including Eyerising (Jiangsu, China; n=14 studies; 650±10 nm, 2.00±0.50 mW),^8–11, 33, 34, 40–42, 44–48^ LD-A (Jilin, China; n=2 studies; 635 nm, 0.35±0.02 mW),^35, 37^ YF020A (Hunan, China; n=3 studies; 650 nm, no specified power),^36, 39, 43^ and sky-n1201 (Beijing, China; n=1 study; 650 nm, with variable outputs of 0.37±0.02 mW, 0.60±0.2 mW, and 1.20 mW).^38^ All studies adhered to a uniform daily RLRL treatment protocol, consisting of two daily 3-minute sessions, with a minimum interval of 4 hours between sessions. Six studies implemented a 5-day per week regimen,^8–10, 41, 43, 46^ while the remaining 14 studies provided treatment 7 days per week.^11, 33–40, 42, 44, 45, 47, 48^

### Safety Profiles of RLRL Therapy

**Table 2** presents the safety outcomes reported among participants undergoing RLRL therapy. In these studies, visual function was assessed using best-corrected visual acuity (BCVA) or multifocal electroretinogram, while ocular structures were evaluated with optical coherence tomography (OCT), fundus photography, or slit-lamp microscopy. Among 529 children assessed for visual function, 528 (99.8%) either maintained a BCVA of ≥20/25 or experienced no decline at follow-up visits compared to baseline. OCT identified no structural damage in 624 (99.8%) of the 625 participants with such data.

**Table 2.**
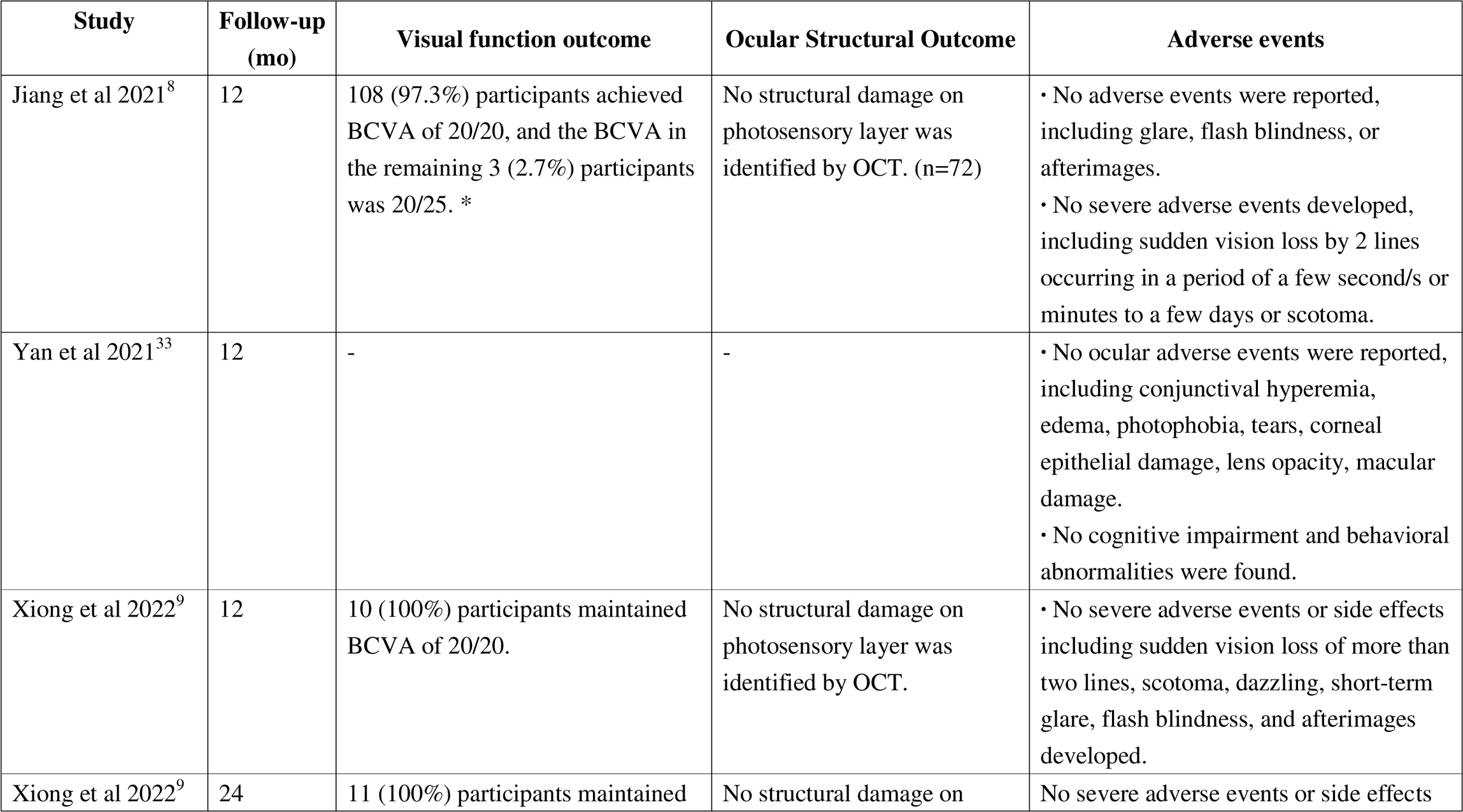
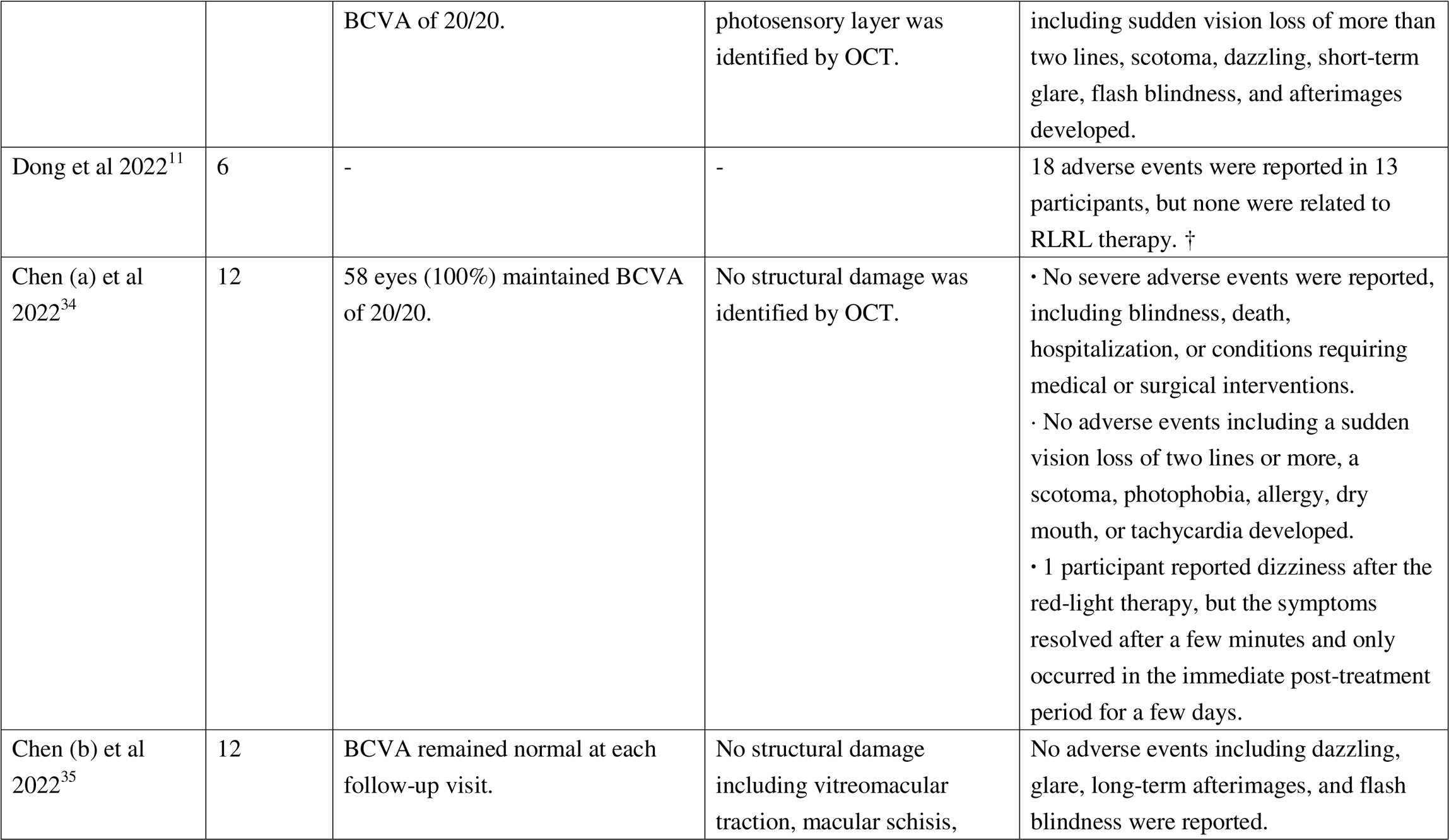
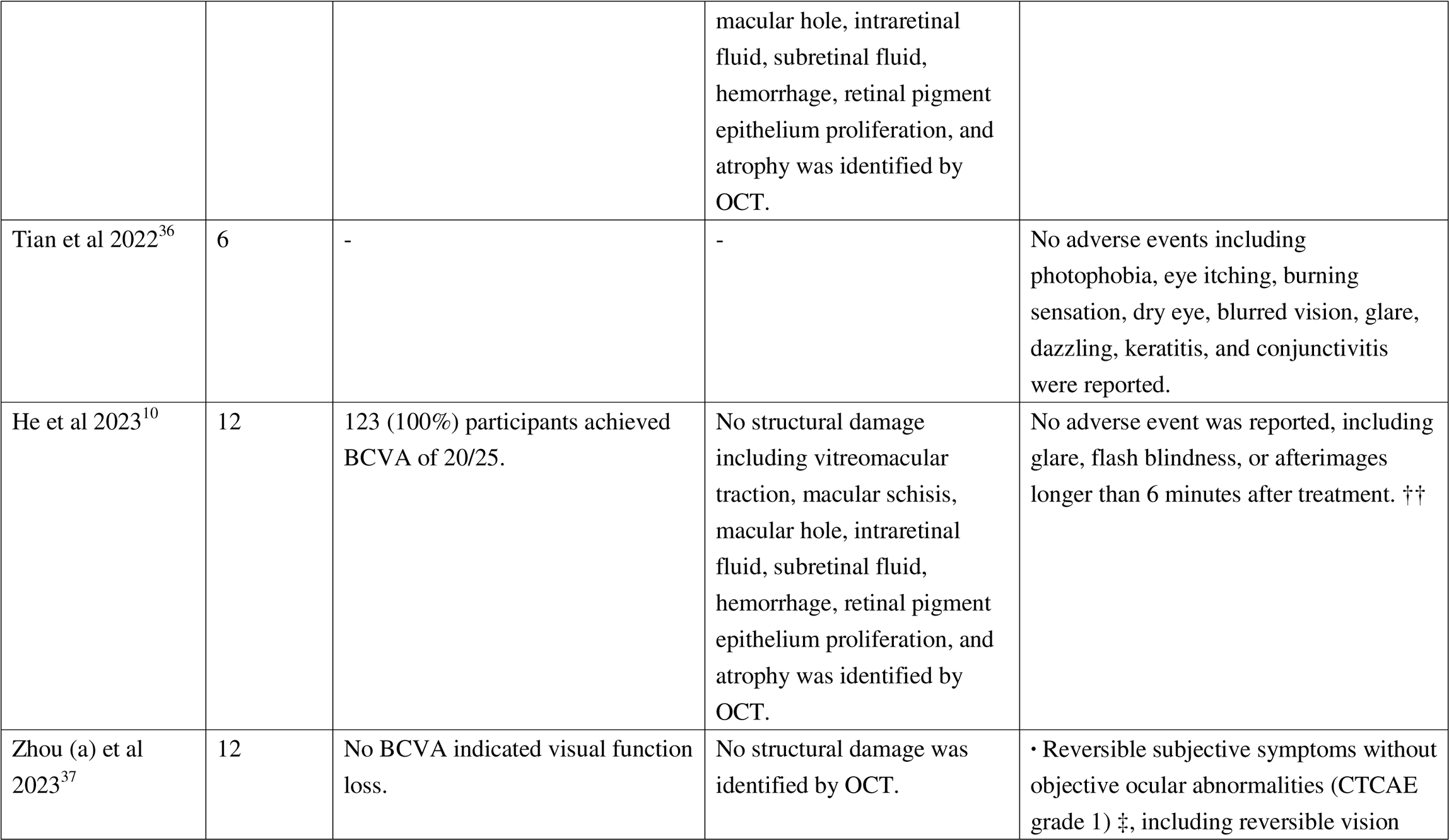
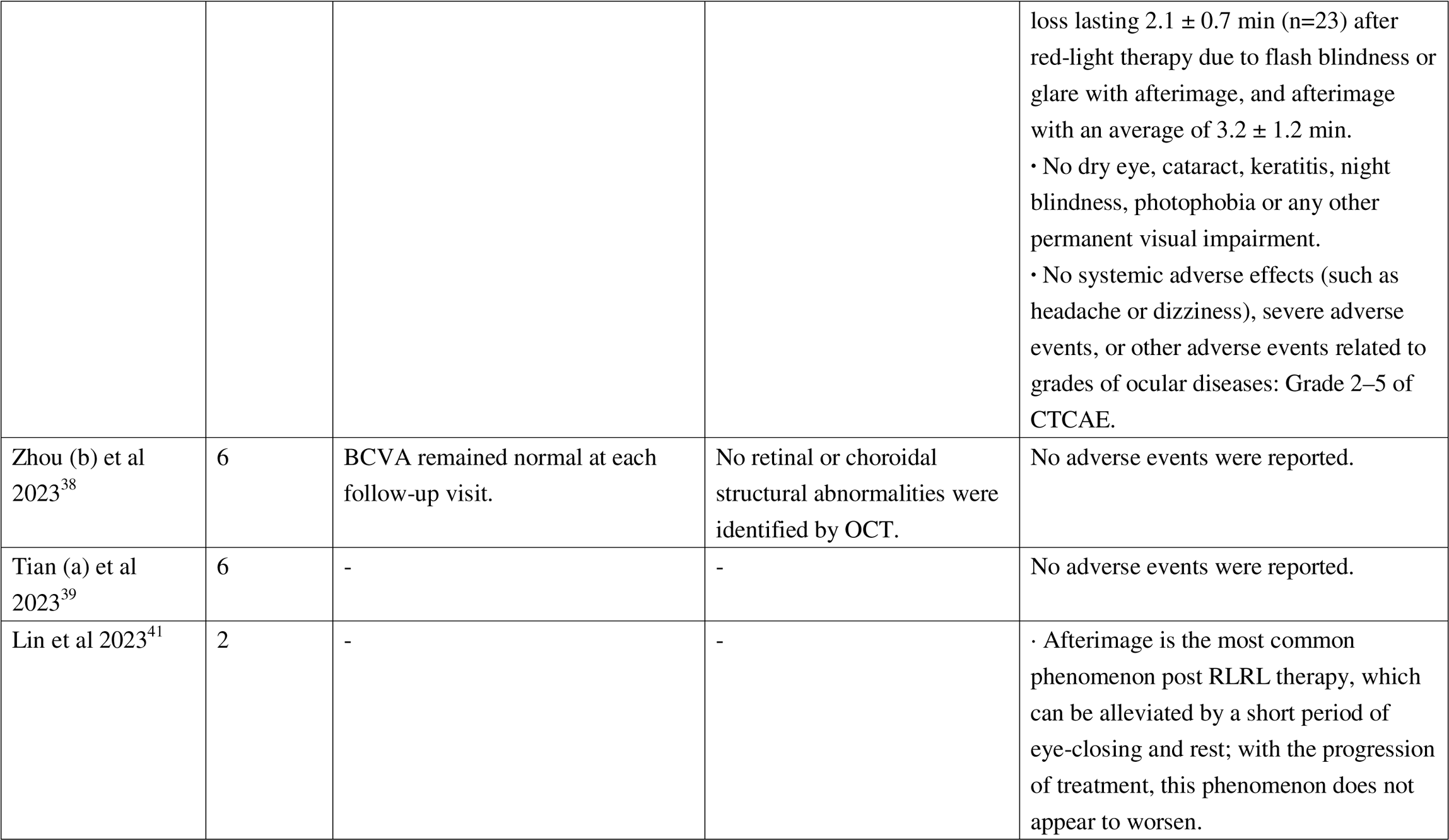
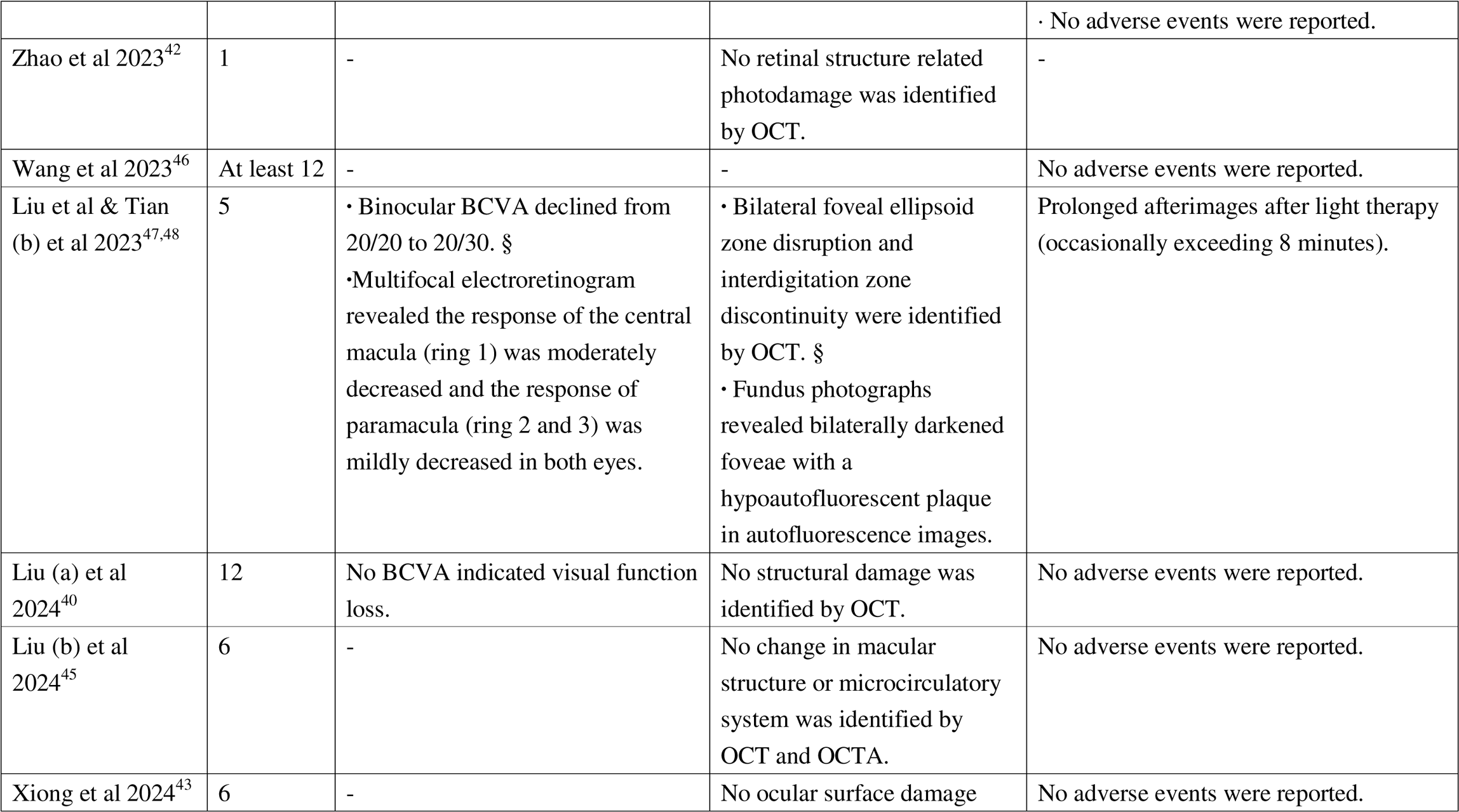
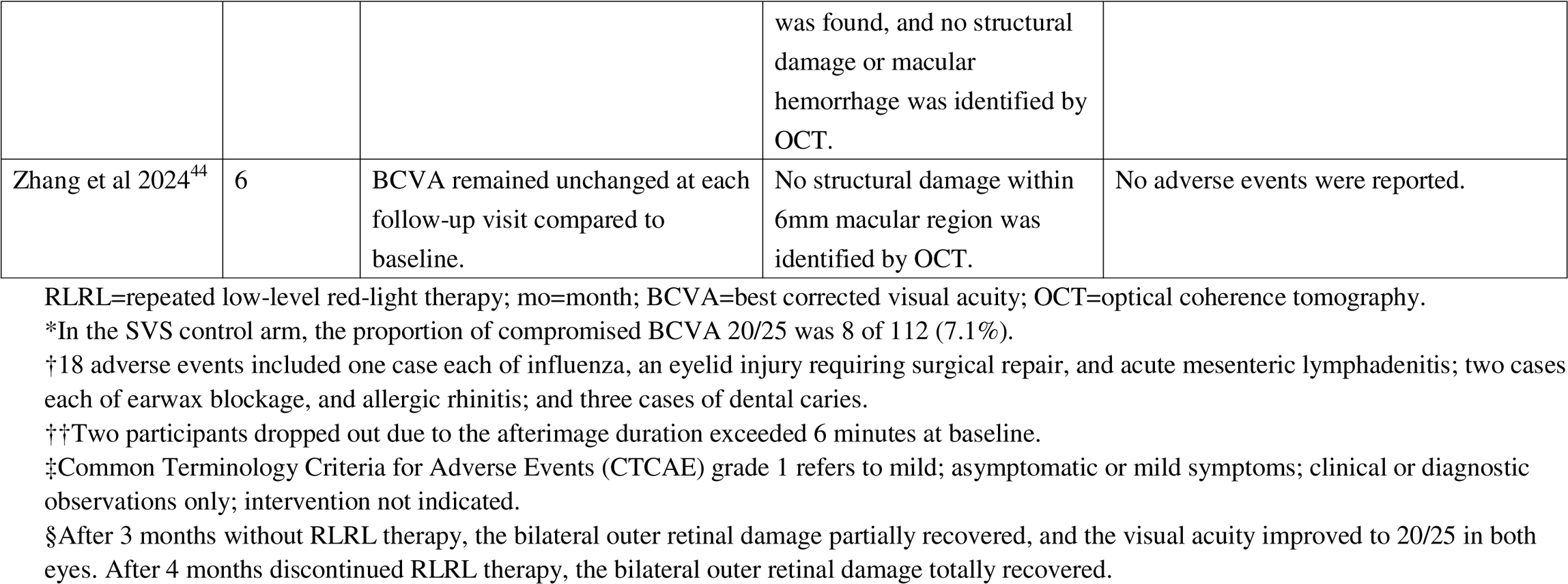
Visual function, ocular structures and adverse events reported in RLRL studies.

Two case reports described the same twelve-year-old girl who experienced a two-line decline in binocular BCVA after five months of RLRL therapy.^47, 48^ The child was highly myopic prior to initiating RLRL, with SER of −6.75D in the right eye and −6.25D in the left eye, and BCVA of 20/25 at the point of initiation. Due to repeated inflammation from orthokeratology, the subject had been switched to RLRL for managing myopia. After three months of RLRL treatment, the BCVA improved to 20/20. After five months for RLRL treatment, the child showed the binocular BCVA decrease from 20/20 to 20/30 with prolonged afterimages occasionally exceeding 8 minutes. Multifocal electroretinogram indicated moderately and mildly decreased response in the macula and paramacular respectively. OCT images showed bilateral disruption of the foveal ellipsoid zone and discontinuity of the interdigitation zone. Fundus photographs revealed bilaterally darkened foveae with a hypoautofluorescent plaque in autofluorescence images. The bilateral outer retinal damage showed partial recovery, with an improvement in binocular BCVA to 20/25, three months after discontinuation of RLRL therapy. Complete recovery of bilateral outer retinal damage was observed four months after discontinuing RLRL therapy.

Nineteen studies reported on ocular adverse events. In one study with 20 participants treated with RLRL, afterimages lasting a mean of 3.2±1.2 minutes were documented without any objective ocular abnormalities.^37^ One trial reported afterimage as the most common phenomenon post light therapy, which can be alleviated by a short period of eye-closing and rest.^41^ Another study demonstrated no afterimages longer than 6 minutes after 1-year RLRL treatment within 126 premyopic children.^10^ One individual experienced dizziness following the red-light therapy; however, this symptom resolved within a few minutes and persisted during the immediate post-treatment period for only a few days.^34^ Aside from the aforementioned case report,^47, 48^ no other included participants experienced vision loss of ≥2 lines, scotoma or treatment-related adverse events post-therapy.

### Risk-to-benefit Analysis

The sole event of temporary vision loss described in the case reports is considered as the numerator,^47, 48^ and the denominator is a conservative estimate of the total number of participants in reported clinical RLRL studies with safety outcomes. The annual incidence of vision loss from RLRL therapy was estimated as 8.77 per 10,000 patient-years. **Table 3** shows the NNH and NNH/NNT estimates for 1 year and 5 years visual impairment associated with RLRL based on Bullimore’s model. A total of 17.5 individuals have to implement RLRL therapy for 5 years to result in 1-year of visual impairment, while 87.7 patients have to use it to result in 5-year of visual impairment.

**Table 3.**
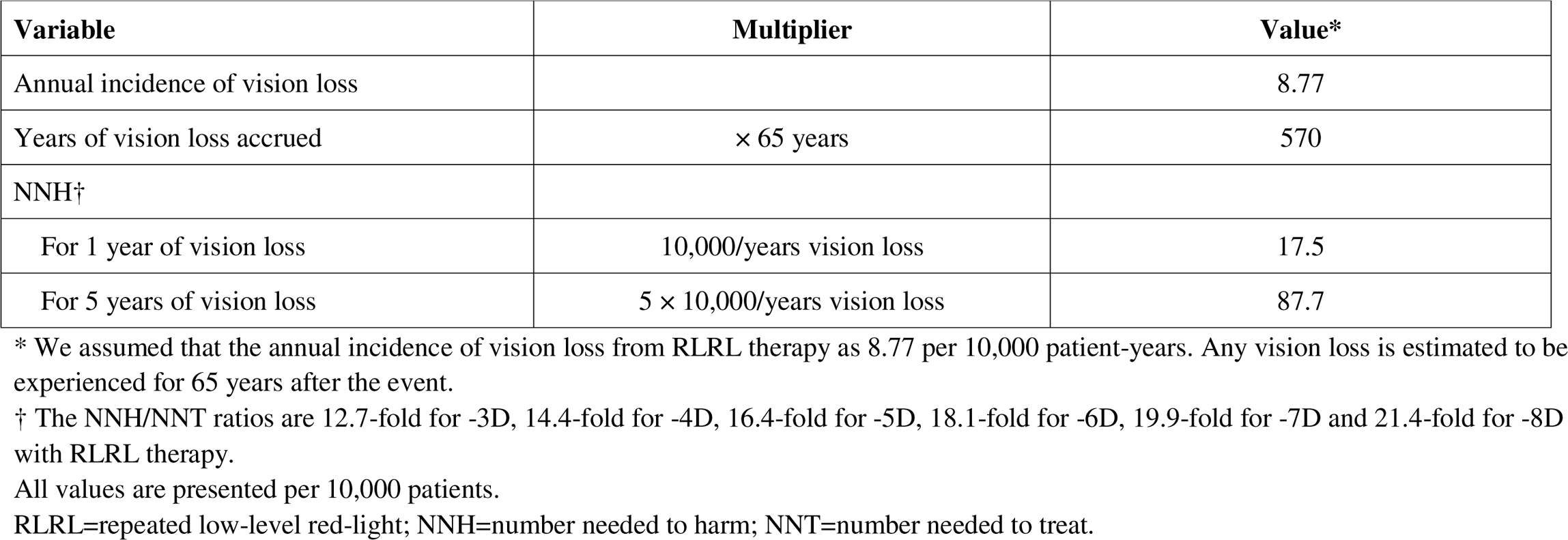
Estimated NNH and NNH/NNT ratios associated with RLRL therapy.

| Variable | Multiplier | Value* |
| --- | --- | --- |
| Annual incidence of vision loss |  | 8.77 |
| Years of vision loss accrued | × 65 years | 570 |
| NNH† |  |  |
| For 1 year of vision loss | 10,000/years vision loss | 17.5 |
| For 5 years of vision loss | 5 × 10,000/years vision loss | 87.7 |

\* We assumed that the annual incidence of vision loss from RLRL therapy as 8.77 per 10,000 patient-years. Any vision loss is estimated to be experienced for 65 years after the event.
† The NNH/NNT ratios are 12.7-fold for -3D, 14.4-fold for -4D, 16.4-fold for -5D, 18.1-fold for -6D, 19.9-fold for -7D and 21.4-fold for -8D with RLRL therapy.
All values are presented per 10,000 patients.
RLRL=repeated low-level red-light; NNH=number needed to harm; NNT=number needed to treat.

From Bullimore’s assessment, controlling myopia by 1D prevents between 0.74 and 1.22 years of visual impairment due to myopic complications across myopia levels of −3D to −8D.^16^ This benefit outweighs the risk of visual impairment years from RLRL treatment, which is 570 per 10,000 patients, or 0.057 year per patient. On the other hand, the NNH outweighs the NNT by a ratio of 12.7-21.4 for a person with −3D to −8D myopia treated with RLRL therapy.

### Comparison with Other Interventions

We further compared the incidence rates of ocular adverse events between RLRL and other myopia interventions. The characteristics and adverse events of other treatments reported in RCTs lasting at least 1 year are displayed in **Table S3**. The side effect incidence of RLRL therapy is 0.088 per 100 patient-years (95% confidence interval [CI], 0.02-0.50), which is comparable to spectacles designed for myopia reduction (0.22 per 100 patient-years; 95% CI, 0.09-0.51; P=0.385),^49–59^ and significantly lower than for low-dose atropine (7.32 per 100 patient-years; 95% CI, 6.65-8.05; P<0.001),^60–78^ orthokeratology (20.6 per 100 patient-years; 95% CI, 16.7-25.0; P<0.001)^77, 79–84^ and other anti-myopia contact lens (19.3 per 100 patient-years; 95% CI, 17.6-21.1; P<0.001)^85–90^ (**Table 4**).

**Table 4.** Comparison of the ocular adverse event incidence rate between RLRL and other interventions.

| **Interventions** | **Baseline age criteria (years)** | **Number of participants** | **Person- years** | **Ocular adverse events** | **Ocular adverse events per 100 patient-years** | **95% CI\*** |
| --- | --- | --- | --- | --- | --- | --- |
| RLRL<sup>8-11,33-48</sup> | 3-18 | 1436 | 1139.8 | 1 | 0.088 | 0.02-0.50 |
| Low-dose atropine<sup>60-78</sup> | 4-16 | 2736 | 5368 | 393 | 7.32 | 6.65-8.05 |
| Orthokeratology<sup>76,79-84</sup> | 6-12 | 231 | 364.5 | 75 | 20.6 | 16.7-25.0 |
| Anti-myopia contact lens<sup>85-90</sup> | 7-15 | 697 | 1899 | 366 | 19.3 | 17.6-21.1 |
| Spectacles designed for myopia reduction<sup>49-59</sup> | 6-16 | 1247 | 2299 | 5 | 0.22 | 0.09-0.51 |

RLRL=repeated low-level red-light; CI=confidence interval.
Anti-myopia contact lens includes rigid gas permeable contact lens and soft multifocal contact lens. Spectacles designed for myopia reduction includes bifocal lens, progressive addition spectacles, aspherical lenslets and peripheral defocus spectacles.
\*95% CI was calculated using Wilson methods.

## Discussion

The results of the systematic review indicated that no visual function loss or ocular structural change with irreversible damage was identified with RLRL therapy. An afterimage was the most common ocular symptom following treatment, with a resolution time of less than 6 minutes reported in two clinical trials. This phenomenon, induced by prior adaptation to a visual stimulus, is believed to be due to the natural bleaching of photochemical pigments or neural adaptation in the retina. Participants with an afterimage duration exceeding 6 minutes were considered clinically too sensitive to the light stimulus.^10, 91^ Among dozens of published studies, only two reported on a single case of a reversible decline in visual function and discontinuity of the foveal ellipsoid and interdigitation zones in an identical girl, with complete recovery after four months of treatment cessation. A risk-to-benefit analysis emphasized that the benefit of reducing visual impairment outweighed the potential risks associated with RLRL treatment. The incidence rate of side effects from RLRL therapy was comparable to that of spectacles wearing, and was lower than that of other therapies.

Of note, two case reports, both describing on complications of the same child following RLRL therapy offers valuable clinical data and serves as a reference point for RLRL’s clinical application.^47, 48^ Given the rarity of this adverse event, it might be secondary to specific individual differences, where the patient may be especially responsive to light therapy. Such individual variability may render the retina more sensitive to light and prone to phototoxicity. It is important to note that the patient was highly myopic, a condition that often involves inherent issues with the retinal structure of the fundus. From the addition information provided by the authors of one case report,^48^ this child experienced significant myopia regression (cycloplegic SER from −6.75D at baseline to −4.50D after 1 month in the right eye; from −6.25D at baseline to −4.50D after 1 month in the left eye) following RLRL treatment. The dark choroid shown in the OCT image post RLRL has been suggested in a non peer-reviewed letter to the editor regarding the case written by experts in inherited retinal disease to represent Stargardt’s disease (https://jamanetwork.com/journals/jamaophthalmology/fullarticle/2805391).

Although the incidence of such adverse events is extremely low, identifying such super-responders to RLRL therapy is still important. From this case, possible characteristics of super-responders include a significant treatment effect, a marked SER regression or AL shortening and an afterimage duration exceeding 6 minutes in response to light exposure. It is crucial for clinicians and parents to closely monitor subjects with these characteristics, as any decrease in visual acuity while wearing glasses may suggest the development of such complications, which can then be further detected and characterized through OCT examination. Importantly, these complications are reversible as after stopping treatment for 4 months, the visual function and ocular structures changes returned to normal. Meticulous supervision is thus necessary throughout the treatment process to ensure safe implementation of RLRL. Appropriate actions include documentation of the retina through fundus photography and OCT before starting treatment and at each routine examination, tracking visual acuity, and recording the duration of any afterimages.^92^

A comprehensive risk-to-benefit assessment of myopia treatments should consider various factors, including the intervention effectiveness in slowing myopia progression, the risk of myopia-related visual impairment, the degree of myopia treated, and the specific risks associated with each intervention. From this risk-to-benefit analysis, the risk of vision loss associated with RLRL treatment is seen to be counterbalanced by its benefits in preventing myopia-related visual impairment with a NNH/NNT ratio of 12.7 to 21.4. Previous studies have reported a NNH/NNT ratio ranging from 5.43 for −3D to 9.15 for −8D for overnight contact lens wearing.^16,93^

Additionally, nearly all interventions for myopia control are associated with some side effects and complications. Our systematic review summarized RCTs with at least one year of follow-up reporting the incidence of side effects from available anti-myopia interventions. Our findings indicate that the incidence of adverse events associated with orthokeratology is 20.6 per 100 patient-years, while it was 19.3 per 100 patient-years for other contact lens. Complications linked to contact lens usage included ocular noninfectious inflammatory events and sight-threatening microbial keratitis.^90, 94–96^ A retrospective study authorized and approved by the US FDA estimated the incidence of microbial keratitis from orthokeratology in children to be 14 per 10,000 patient-years (95% CI, 1.7-50.4 per 10,000 patient-years).^97^ Atropine, even at low concentrations of 0.01%, may lead to pupil dilation and loss of accommodation, with photophobia, reduced near vision, and allergic conjunctivitis being commonly reported ocular side effects.^98, 99^ Spectacles in comparison are a more well-tolerated method for correcting and controlling myopia; however, they are associated with a low risk of falls and bicycle collisions.^51, 100^ In this review, RLRL therapy had a comparable incidence of side effects with spectacles.

This systematic review provides the first comprehensive evidence on the safety profile and the risk-to-benefit ratio of RLRL therapy for myopia control. However, the results should be interpreted within the context of several limitations. Firstly, most studies lasted for 12 months, with only 11 children undergoing 24-month treatment in one post-trial study included thereby limiting long-term evidence. There is a need for further large-scale studies to thoroughly assess the long-term safety of RLRL therapy in children and adolescents. Secondly, visual function in the included studies was primarily evaluated using visual acuity, which could be influenced by subjective factors. Objective assessments, such as multifocal electroretinography or microperimetry, are necessary to provide more comprehensive safety evidence for RLRL therapy. Thirdly, the methodology for reporting adverse events varied widely across studies. Some studies documented the number of patients experiencing side effects, whereas others detailed the number of adverse events. Fourthly, the accuracy of Bullimore’s model is dependent on the validity of its assumptions.^16^ It presumses that the risk of vision loss from RLRL therapy is independent of refractive error. The model also assumes a fixed treatment effect with myopia control. Fifthly, quality appraisal was employed exclusively for the included RCTs, as the Cochrane Risk of Bias Tool is not designed for use with non-randomized controlled trial, post-trial study, retrospective study, single-arm study or case report. Finally, eleven of the twenty studies were RCTs, which were conducted under strict surveillance. Such studies may not accurately reflect real-world vision care, therefore more real-world studies are needed to better understand long-term safety of RLRL in a greater variety of settings.

## Conclusion

In conclusion, no irreversible visual function loss or ocular structural damage associated with RLRL therapy was identified in this review. Meticulous supervision is crucial throughout the entire treatment process by clinicians and parents, including documenting the status of retina through fundus photography and OCT before initiatng RLRL therapy and at each routine examination, as well as tracking visual acuity and the duration of any afterimages at home. Screening for and early identification of rare super-responders is also important to avoid potential light injury. Future larger and longer-term real-world studies are needed to better understand the long-term safety of RLRL.

## Data Availability

All data produced in the present work are contained in the manuscript.

## Abbreviations and Acronyms

RLRL: repeated low-level red-light
OCT: optical coherence tomography
CI: confidence interval
IMI: International Myopia Institute
RCT: randomized controlled trial
SER: spherical equivalent refraction
AL: axial length
NNH: number needed to harm
NNT: number needed to treat
SVS: single-vision spectacles
BCVA: best-corrected visual acuity.

## Acknowledgment

The authors thank the Hainan Province Clinical Medical Center, the National Natural Science Foundation of China (82371086) for their support.

## Author Contributions

Prof. Wang had full access to all the data in the study and took responsibility for the integrity of the data and the accuracy of the data analysis.

Study concept and design: Chen, Congdon, Wang.

Data collection: Chen, Chen, Xiong, Yang, Wang.

Analysis and interpretation of data: Chen, Wang.

Drafting of the manuscript: Chen, Congdon, Wang.

Critical revision of the manuscript for important intellectual content: All authors.

Statistical analysis: Chen, Wang.

Obtained funding: Wang.

Administrative, technical, or material support: Congdon, Wang.

Overall supervision: Wang.

