## Supplemental Materials for "Safety of Repeated Low-Level Red-Light Therapy for Myopia: A Systematic Review"

**Supplementary Table 1.** Search strategy (PubMed style).

| **Concept** | **Search string** |
| --- | --- |
| #1 - Myopia | (myopia[MeSH]) OR (myopia[Title/Abstract]) OR (myop*[Title/Abstract]) OR (short-sightedness[Title/Abstract]) OR (short sight[Title/Abstract]) OR (short sighted[Title/Abstract]) OR (near-sightedness[Title/Abstract]) OR (near sight[Title/Abstract]) OR (near sighted[Title/Abstract]) OR (refractive errors[Title/Abstract]) |
| #2 - RLRL | (Low-Level Light Therapy[MeSH]) OR (Low-Level Light Therapy[Title/Abstract]) OR (repeated low-level red-light therapy[Title/Abstract]) OR (repeated red-light therapy[Title/Abstract]) OR (repeated low-intensity red light therapy[Title/Abstract]) OR (low power laser therapy[Title/Abstract]) OR (red light [Title/Abstract]) OR (RLRL[Title/Abstract]) OR (LLLT[Title/Abstract]) OR (photobiomodulation[Title/Abstract]) |
| #3 | #1 AND #2 |

**Supplementary Table 2**. Specifications of RLRL device and treatment regimens adopted in RLRL studies.

| **Study** | **Device name and manufacture** | **Wavelength (mm)** | **Power (mW)** | **Regimen** |
| --- | --- | --- | --- | --- |
| Jiang et al 2021^8^ | Eyerising; Suzhou Xuanjia Optoelectronics Technology, Jiangsu, China | 650 ± 10 | 2.00 ± 0.50 | 5 days per week  3 mins per session, twice daily with a minimum interval of 4 hours |
| Yan et al 2021^33^ | Eyerising; Suzhou Xuanjia Optoelectronics Technology, Jiangsu, China | 650 ± 10 | 2.00 ± 0.50 | 7 days per week  3 mins per session, twice daily with a minimum interval of 4 hours |
| Xiong et al 2022^9^ | Eyerising; Suzhou Xuanjia Optoelectronics Technology, Jiangsu, China | 650 ± 10 | 2.00 ± 0.50 | 5 days per week  3 mins per session, twice daily with a minimum interval of 4 hours |
| Dong et al 2022^11^ | Eyerising; Suzhou Xuanjia Optoelectronics Technology, Jiangsu, China | 650 ± 10 | 2.00 ± 0.50 | 7 days per week  3 mins per session, twice daily with a minimum interval of 4 hours |
| Chen (a) et al 2022^34^ | Eyerising; Suzhou Xuanjia Optoelectronics Technology, Jiangsu, China | 650 ± 10 | 2.00 ± 0.50 | 7 days per week  3 mins per session, twice daily with a minimum interval of 4 hours |
| Chen (b) et al 2022^35^ | LD-A, Jilin Londa Optoelectronics Technology, Jilin, China | 635 | 0.35 ± 0.02 | 7 days per week  3 mins per session, twice daily with a minimum interval of 4 hours |
| Tian et al 2022^36^ | YF020A; Hunan EnVan Technology Co.,Ltd., Hunan, China | 650 | - | 7 days per week  3 mins per session, twice daily with a minimum interval of 4 hours |
| He et al 2023^10^ | Eyerising; Suzhou Xuanjia Optoelectronics Technology, Jiangsu, China | 650 ± 10 | 2.00 ± 0.50 | 5 days per week  3 mins per session, twice daily with a minimum interval of 4 hours |
| Zhou (a) et al 2023^37^ | LD-A, Jilin Londa Optoelectronics Technology, Jilin, China | 650 ± 10 | 0.35 ± 0.02 | 7 days per week  3 mins per session, twice daily with a minimum interval of 4 hours |
| Zhou (b) et al 2023^38^ | sky-n1201; Beijing Ming Ren Shi Kang Science & Technology Co., Ltd., Beijing, China | 650 | 0.37 ± 0.02; 0.60 ± 0.2;  1.20 | 7 days per week  3 mins per session, twice daily with a minimum interval of 4 hours |
| Tian (a) et al 2023^39^ | YF020A; Hunan EnVan Technology Co.,Ltd., Hunan, China | 650 | - | 7 days per week  3 mins per session, twice daily with a minimum interval of 4 hours |
| Lin et al 2023^41^ | Eyerising; Suzhou Xuanjia Optoelectronics Technology, Jiangsu, China | 650 ± 10 | 2.00 ± 0.50 | 5 days per week  3 mins per session, twice daily with a minimum interval of 4 hours |
| Zhao et al 2023^42^ | Eyerising; Suzhou Xuanjia Optoelectronics Technology, Jiangsu, China | 650 ± 10 | 2.00 ± 0.50 | 7 days per week  3 mins per session, twice daily with a minimum interval of 4 hours |
| Wang et al 2023^46^ | Eyerising; Suzhou Xuanjia Optoelectronics Technology, Jiangsu, China | 650 ± 10 | 2.00 ± 0.50 | 5 days per week  3 mins per session, twice daily with a minimum interval of 4 hours |
| Liu et al & Tian (b) et al 2023^47,48^ | Eyerising; Suzhou Xuanjia Optoelectronics Technology, Jiangsu, China | 650 ± 10 | 2.00 ± 0.50 | 7 days per week  3 mins per session, twice daily with a minimum interval of 4 hours |
| Liu (a) et al 2024^40^ | Eyerising; Suzhou Xuanjia Optoelectronics Technology, Jiangsu, China | 650 ± 10 | 2.00 ± 0.50 | 7 days per week  3 mins per session, twice daily with a minimum interval of 4 hours |
| Liu (b) 2024^45^ | Eyerising; Suzhou Xuanjia Optoelectronics Technology, Jiangsu, China | 650 ± 10 | 2.00 ± 0.50 | 7 days per week  3 mins per session, twice daily with a minimum interval of 4 hours |
| Xiong et al 2024^43^ | YF020A; Hunan Yifan Technology Co., LTD., Hunan, China | 650 | - | 5 days per week  3 mins per session, twice daily with a minimum interval of 4 hours |
| Zhang et al 2024^44^ | Eyerising; Suzhou Xuanjia Optoelectronics Technology, Jiangsu, China | 650 ± 10 | 2.00 ± 0.50 | 7 days per week  3 mins per session, twice daily with a minimum interval of 4 hours |

RLRL=repeated low-level red-light.

**Supplementary Table 3**. Characteristics and adverse events of other interventions reported in randomized controlled trials lasting at least 1 year.

| **Author Year (Country)** | **Follow-up (mo)** | **Age** | **No. of patients** | **Intervention and control** | **Adverse events** |
| --- | --- | --- | --- | --- | --- |
| **Low-dose atropine** | | | | | |
| Yen et al 1989 (China, Taiwan)^60^ | 12 | 6-14 | 32 | 1% Atropine | Photophobia: 32 cases  No systemic or ocular complications were observed. |
|  |  |  | 32 | 1% Cyclopentolate | No systemic or ocular complications were observed. |
|  |  |  | 32 | Placebo eyedrops | No systemic or ocular complications were observed. |
| Shin et al 1999 (China, Taiwan)^61^ | 24 | 6-13 | 41 | 0.5% Atropine | Photophobia: 5 cases Recurrent allergic blepharitis: 1 case |
|  |  |  | 47 | 0.25% Atropine | Photophobia/near work problems: 3 cases No systemic or ocular complications were observed. |
|  |  |  | 49 | 0.1% Atropine | No systemic or ocular complications were observed. |
|  |  |  | 49 | Tropicamide | No systemic or ocular complications were observed. |
| Chua et al 2006 (Singapore)^62^ | 24 | 6-12 | 166 | 1% Atropine | No serious adverse events related to atropine were reported. |
|  |  |  | 190 | Vehicle eye drops | No serious adverse events were reported. |
| Chia et al 2012 (Singapore)^63^ | 24 | 6-12 | 161 | 0.5% Atropine | Allergic conjunctivitis: 7 cases  Allergy-related dermatitis of the eyelids: 4 cases  Chalazion: 16 cases  Loss of BCVA>1 line:13 cases  Intolerable glare: 1  No severe adverse events related to atropine treatment. |
|  |  |  | 155 | 0.1% Atropine | Allergic conjunctivitis: 7 cases Allergy-related dermatitis of the eyelids: 2 cases Chalazion: 16 cases Loss of BCVA>1 line: 20 cases No severe adverse events related to atropine treatment. |
|  |  |  | 84 | 0.01% Atropine | Chalazion: 2 cases  Loss of BCVA>1 line: 11 cases  Irritation: 1  Blur: 1  No severe adverse events related to atropine treatment. |
| Yi et al 2015 (China, Taiwan)^64^ | 12 | 7-12 | 62 | 1% Atropine | No adverse events were observed. |
|  |  |  | 62 | Placebo eyedrops | No adverse events were observed. |
| Wang et al 2017 (China)^65^ | 12 | 9.1 (1.4) | 54 | 0.5% Atropine | No serious adverse events were observed, such as eyes itching and distention. |
|  |  | 8.7 (1.5) | 55 | Placebo eyedrops | No serious adverse events were observed, such as eyes itching and distention. |
| Yam et al 2019 (China, Hongkong)^66^ | 12 | 4-12 | 109 | 0.05% Atropine | Photophobia at 1 year: 8 cases Allergic conjunctivitis: 3 cases |
|  |  |  | 108 | 0.025% Atropine | Photophobia at 1 year: 6 children Allergic conjunctivitis: 7 children |
|  |  |  | 110 | 0.01% Atropine | Photophobia at 1 year: 2 cases Allergic conjunctivitis: 7 cases |
|  |  |  | 111 | Placebo eyedrops | Photophobia at 1 year: 4 cases Allergic conjunctivitis: 7 cases |
| Wei et al 2020 (China)^67^ | 12 | 6-12 | 76 | 0.01% Atropine | Photophobia: 5 cases Allergic conjunctivitis: 4 cases No serious adverse events associated with atropine were reported. |
|  |  |  | 83 | Placebo eyedrops | Photophobia: 1 case Allergic conjunctivitis: 1 case No serious adverse events were reported. |
| Yam et al 2020 (China, Hongkong)^68^ | 24 | 4-12 | 93 | 0.05% Atropine | Photophobia: 8 cases Allergic conjunctivitis: 9 cases |
|  |  |  | 86 | 0.025% Atropine | Photophobia: 4 cases Allergic conjunctivitis: 10 cases |
|  |  |  | 91 | 0.01% Atropine | Photophobia: 6 cases Allergic conjunctivitis: 11 cases |
|  | 12 | 4-12 | 80 | Switch to 0.5% Atropine | Photophobia: 7 cases Allergic conjunctivitis: 7 cases |
| Cui et al 2021 (China)^69^ | 24 | 6-14 | 105 | 0.02% Atropine | Photophobia in bright sunlight: 32 cases |
|  |  |  | 106 | 0.01% Atropine | Photophobia in bright sunlight: 32 cases |
|  |  |  | 89 | SVS | Photophobia in bright sunlight: 3 cases |
| Hieda et al 2021 (Japan)^70^ | 24 | 6-12 | 84 | 0.01% Atropine | Photophobia: 1 case Allergic conjunctivitis: 2 cases |
|  |  |  | 84 | Placebo eyedrops | Near-vision impairment: 1 case  Allergic conjunctivitis: 1 cases |
| Saxena et al 2021 (India)^71^ | 12 | 6-14 | 47 | 0.01% Atropine | No blurring or photophobia was observed. |
|  |  |  | 45 | Placebo eyedrops | No blurring or photophobia was observed. |
| Lee et al 2022 (Australia)^72^ | 24 | 6-16 | 94 | 0.01% Atropine | Sore or heavy-feeling eye: 2 cases Blurred near vision: 1 case |
|  |  |  | 37 | Placebo eyedrops | No adverse events related to treatment were reported. |
| Yam et al 2022 (China, Hongkong)^73^ | 36 | 4-12 | 45 | 0.05% Atropine | Photophobia: 1 case Allergic conjunctivitis: 2 cases No serious adverse events associated with atropine were reported. |
|  |  |  | 39 | 0.025% Atropine | Photophobia: 3 cases Allergic conjunctivitis: 2 cases No serious adverse events associated with atropine were reported. |
|  |  |  | 43 | 0.01% Atropine | Photophobia: 2 cases Allergic conjunctivitis: 2 cases No serious adverse events associated with atropine were reported. |
| Chia et al 2023 (Singapore)^74^ | 12 | 6-11 | 24 | 0.0025% Atropine | Vision blurred: 2 cases Eye pruritus: 2 cases Glare: 1 case  Eye pain: 2 cases  Eye infection: 1 case No serious adverse events associated with atropine were reported. |
|  |  |  | 24 | 0.005% Atropine | Vision blurred: 1 case Eye swelling: 1 case No serious adverse events associated with atropine were reported. |
|  |  |  | 25 | 0.01% Atropine | Vision blurred: 1 case Allergic conjunctivitis: 1 case Eye pruritus: 2 cases Glare: 2 cases  Ocular hyperemia: 1 case No serious adverse events associated with atropine were reported. |
|  |  |  | 26 | Placebo eyedrops | No ocular adverse events were reported. |
| Hansen et al 2023 (Denmark)^75^ | 12 | 6-12 | 32 | 0.01% Atropine | Blurred near vision: 1 case |
|  |  |  | 29 | Placebo eyedrops | Blurred near vision: 1 case |
| Moriche-Carretero et al 2023 (Spain)^76^ | 60 | 6.8 (1.9) | 184 | 0.01% Atropine | No side effects were reported. |
|  |  | 6.7 (1.9) | 177 | Blank control | No side effects were reported. |
| Xu et al 2023 (China)^77^ | 24 | 8-12 | 42 | 0.01% Atropine | Photophobia: 2 cases |
|  |  |  | 40 | OK | Suspected superficial punctuate keratitis: 2 cases |
|  |  |  | 42 | 0.01% Atropine+OK | Mechanical corneal abrasion: 1 case  Photophobia: 1 case |
|  |  |  | 40 | Placebo eyedrops | No ocular adverse events were reported. |
| Yam et al 2023 (China, Hongkong)^78^ | 24 | 4-9 | 116 | 0.05% Atropine | Photophobia: 15 cases allergic conjunctivitis: 5 cases |
|  |  |  | 122 | 0.01% Atropine | Photophobia: 23 cases  Allergic conjunctivitis: 3 cases |
|  |  |  | 125 | Placebo eyedrops | Photophobia: 14 cases Allergic conjunctivitis: 2 cases |
| **OK lens** | | | | | |
| Cho et al 2012 (China, Hongkong)^79^ | 24 | 6-10 | 37 | OK | Corneal staining: 3 cases Conjunctival hyperemia:1 case Chalazion: 1 case |
|  |  |  | 41 | SVS | Recurrent corneal inflammation: 1 case |
| Charm et al 2013 (China, Hongkong)^80^ | 24 | 8-11 | 12 | OK | Corneal staining (Grade 1): 5 cases Pigmented arc: 12 cases |
|  |  |  | 16 | SVS | Corneal staining (Grade 1): 5 cases |
| Kinoshita et al 2020 (Japan)^81^ | 24 | 8-12 | 35 | OK | Corneal infiltrates: 1 case Mild superficial punctate keratitis: 1 case |
|  |  |  | 38 | OK+0.01% Atropine | Mild superficial punctate keratitis: 2 cases |
| Tan et al 2020 (China, Hongkong)^82^ | 12 | 6-11 | 30 | OK | Infiltrative keratitis: 1 case Bacterial conjunctivitis: 1 case |
|  |  |  | 29 | OK+0.01% Atropine | Bacterial conjunctivitis: 1 case |
| Guo et al 2021 (China, Hongkong)^83^ | 12 | 6-11 | 32 | OK-type I | Lens binding: 12 cases Corneal staining: 11 cases Infiltrate: 3 cases Microcysts (Grade 2): 1 case  Corneal neovascularization: 1 case |
|  |  |  | 26 | OK-type II | Lens binding: 7 cases Corneal staining: 6 cases Infiltrate: 3 cases |
| Jakobsen et al 2022 (Denmark)^84^ | 18 | 6-12 | 19 | OK | Corneal staining: 3 cases (each case of Grade 1, 2 and 3) |
|  |  |  | 28 | SVS | No adverse events were reported. |
| Xu et al 2023 (China)^77^ | 24 | 8-12 | 42 | 0.01% Atropine | Photophobia: 2 cases |
|  |  |  | 40 | OK | Suspected superficial punctuate keratitis: 2 cases |
|  |  |  | 42 | 0.01% Atropine+OK | Mechanical corneal abrasion: 1 case  Photophobia: 1 case |
|  |  |  | 40 | Placebo eyedrops | No ocular adverse events were reported. |
| **Contact lens** | | | | | |
| Walline et al 2004 (USA)^85^ | 36 | 8-11 | 59 | RGP | No sight-threatening adverse event |
|  |  |  | 57 | SCL | Allergy: 1 case  3 adverse events due to tight-fitting SCLs |
| Cheng et al 2016 (USA)^86^ | 12 | 8-11 | 64 | Positive spherical aberrtion SCL | Allergic conjunctivitis: 1 cases |
|  |  |  | 63 | SCL | Contact dermatitis: 1 cases Corneal neovascularization (Grade 2): 1 case |
| Chamberlain et al 2019 (USA)^87^ | 36 | 8-12 | 53 | MiSight 1-day | 7 cases of corneal infiltrative, foreign body, bilateral allergic reaction, unilateral mild pannus (requiring temporary discontinuation), superficial punctate corneal staining, a unilateral subconjunctival hemorrhage, and a case of irritation with the lens located under the eyelid.  Palpebral roughness (Grade 3): 1 case |
|  |  |  | 56 | Proclear 1-day | 7 cases of corneal infiltrative, foreign body, bilateral allergic reaction, unilateral mild pannus (requiring temporary discontinuation), superficial punctate corneal staining, a unilateral subconjunctival hemorrhage, and a case of irritation with the lens located under the eyelid. |
| Walline et al 2020 (USA)^88^ | 36 | 7-11 | 97 | High add power (+2.50D) SCL | Giant papillary conjunctivitis:9 cases Infiltrative keratitis: 8 cases Allergy: 7 cases Corneal epithelial defect:3 cases Contact lens associated red eye: 2 cases Sterile corneal ulcer: 2 cases Toxic in eye: 2 cases Superior epithelial arcuate lesion: 1 case Probable microbial keratitis: 1 case |
|  |  |  | 98 | Medium add power (+1.50D) SCL |  |
|  |  |  | 97 | SCL |  |
| Valle et al 2020 (Spain)^89^ | 12 | 7-15 | 32 | Progressive multifocal and reverse geometry SCL (Esencia) | Conjunctival hyperaemia: 1 case Corneal neovascularisation: 3 cases Micropapillary response: 3 cases Papillary conjunctivitis: 1 case Superficial punctuate keratitis: 2 cases |
|  |  |  | 26 | SCL | Corneal neovascularisation: 1 case Micropapillary response: 2 cases Superficial punctuate keratitis: 1 case |
| Giannoni et al 2022 (USA)^90^ | 36 | 7-11 | 294 | SCL, multifocal +1.50D SCL, multifocal +2.50D SCL | Adnexa events: 28 cases Corneal events: 153 cases Conjunctiva events: 34 cases  Posterior segment events: 13 cases  Other ocular events: 97 cases |
| **Spectacles** | | | | | |
| Gwiazda et al 2003 (USA)^49^ | 36 | 6-11 | 229 | PAL +2.00D | No serious adverse events were reported. |
|  |  |  | 233 | SVS | No serious adverse events were reported. |
| Cheng et al 2010 (Australia)^50^ | 24 | 8-13 | 48 | Bifocal lens | No adverse events were reported. |
|  |  |  | 46 | Prismatic bifocal lens | No adverse events were reported. |
|  |  |  | 41 | SVS | No adverse events were reported. |
| Sankaridurg et al 2010 (Australia)^51^ | 12 | 6-16 | 48 | Type I novel spectacles | Blurred side vision: 1 case |
|  |  |  | 58 | Type II novel spectacles | Fall when running: 2 cases |
|  |  |  | 46 | Type III novel spectacles | No adverse events were reported. |
|  |  |  | 49 | SVS | No adverse events were reported. |
| COMET2 Group 2011^52^ | 36 | 8-12 | 52 | PAL +2.00D | Distance blur:1 case |
|  |  |  | 58 | SVS | Reduced visual acuity: 1 case Eye pain and change in distance correction: 1 case |
| Hasebe et al 2014 (Japan)^53^ | 24 | 6-12 | 58 | PA-PAL +1.0D | No serious adverse events were reported. |
|  |  |  | 51 | PA-PAL +1.5D | No serious adverse events were reported. |
|  |  |  | 60 | SVS | No serious adverse events were reported. |
| Kanda et al 2018 (Japan)^54^ | 24 | 6-12 | 101 | PAL +1.9D | No adverse events were reported. |
|  |  |  | 102 | SVS | No adverse events were reported. |
| Lam et al 2018 (China, Hongkong)^55^ | 24 | 8-13 | 79 | DIMS | No treatment-related adverse event was reported. |
|  |  |  | 81 | SVS | No treatment-related adverse event was reported. |
| Bao et al 2021 (China)^56^ | 12 | 8-13 | 54 | HAL | No treatment-related adverse event was reported. |
|  |  |  | 55 | SAL | No treatment-related adverse event was reported. |
|  |  |  | 52 | SVS | No treatment-related adverse event was reported. |
| Bao et al 2022 (China)^57^ | 24 | 8-13 | 54 | HAL | No serious adverse events were reported. |
|  |  |  | 53 | SAL | Hit by a ball: 1 case |
|  |  |  | 50 | SVS | Punctate corneal epithelial defects: 1 case |
| Liu et al 2023 (China)^58^ | 12 | 8-12 | 52 | Cylindrical annular refractive element (CARE) | No adverse events were reported |
|  |  |  | 44 | SVS | No adverse events were reported |
| Rappon et al 2023 (USA)^59^ | 12 | 6-10 | 88 | Diffusion optics technology (DOT 0.2) | No treatment-related adverse event was reported. |
|  |  |  | 75 | Diffusion optics technology (DOT 0.24) | No treatment-related adverse event was reported. |
|  |  |  | 93 | SVS | No treatment-related adverse event was reported. |

OK=orthokeratology; SVS=single vision spectacles; SA=spherical aberration; RGP= rigid gas-permeable; SCL=soft contact lens; PA-PAL=positively aspherized progressive addition lenses; DIMS=defocus incorporated multiple segments; HAL=highly aspherical lenslets; SAL=slightly aspherical lenslets; mo=month.
